# Early-life socioeconomic circumstances and the comorbidity of depression and overweight in adolescence and young adulthood: a longitudinal study

**DOI:** 10.1101/2023.04.24.23289020

**Authors:** Fanny Kilpi, Laura D Howe

## Abstract

**Background:** Depression and overweight both often emerge early in life and have been found to be associated, but few studies examine depression-overweight comorbidity and its social patterning early in the life course. This study investigates how different aspects of early-life socioeconomic circumstances are associated with depression-overweight comorbidity from adolescence to young adulthood exploring any differences by age and sex.

**Methods:** Drawing on data from 4,948 participants of the Avon Longitudinal Study of Parents and Children (ALSPAC) birth cohort from the UK, we estimated how parental education, social class and financial difficulties reported in pregnancy were associated with depression and overweight, and their comorbidity at approximately the ages 17 and 24 in males and females.

**Results:** The results from multinomial logistic regression models showed that all three socioeconomic markers were associated with depression-overweight comorbidity and results were similar across age. Lower parental education (relative risk ratio (RRR) and 95% confidence interval (CI) of low education v high education: 3.61 (2.30-5.67) in females and 1.54 (1.14-2.07) in males) and social class (class IV/I v class I: 5.67 (2.48-12.94) in females and 3.11 (0.70-13.91) in males) had strong associations with comorbidity at age 17 relative to having neither depression or overweight. Financial difficulties were also a risk factor in females, with less clear results in males.

**Conclusion:** The findings indicate that early socioeconomic circumstances are linked with the accumulation of mental and physical health problems already in adolescence, which has implications for life-long health inequalities.

## INTRODUCTION

Overweight or obesity and depression in adolescence and young adulthood are important public health concerns as both tend to track into later life (1–3). Overweight and depression may share risk factors and aetiology (4), and studies have also consistently demonstrated a reciprocal relationship between overweight and depression that may begin already at a young age (5–12). The comorbidity of overweight and depression is likely to be one of the earliest common life course manifestations of multimorbidity, with potentially serious and lifelong implications for subsequent chronic diseases and healthcare costs. However, their comorbidity in adolescence and young adulthood has been seldom studied, with research to date focusing on the accumulation of health problems in later life (13). Nevertheless, this period is formative in the development of physical and mental health and health-related behaviours, and an important life stage during which influences from the family translate to health.

Family socioeconomic circumstances have been found to be associated with higher adolescent depressive symptoms (14–17) and BMI (18–20). Disadvantaged circumstances may increase individuals ‘exposure to stress and ability to access resources that help cope with stress. The socioeconomic position (SEP) of the family in general reflects the life chances available through money, power, knowledge and prestige (21). However, parental education, social class and financial difficulties reflect access to somewhat different resources and exposure to different patterns of risk factors, which may not be associated with adolescent health exactly the same way (19,22–24). Education, for example, may influence BMI through access to knowledge about healthier food choices and capacity to adhere to a healthier diet or take advantage of opportunities for physical activity (25). Experiencing financial difficulties can expose to stress that directly affects mental health and can impact sense of personal control. Understanding the social determinants of comorbidity can inform public health measures and improve our understanding of the impact of early-life exposures on potentially accumulative health disadvantage.

Sex and/or gender may influence the relationship between social factors and mental and physical health comorbidity. Prior studies of sex differences in the effects of family SEP on BMI (19,20) or mental health problems (16,17) have returned inconsistent findings. In general, females report higher levels of emotional problems, and an earlier and higher incidence of depression than males (26). At the same time, the relationship between depression and overweight appears to be stronger in females (4,11,27), potentially due to the gendered nature of weight stigma (28).

We examine how parental education, social class and financial difficulties are associated with the comorbidity of depression and overweight in adolescence and early adulthood. These two time-points represent before and after a key life course transition, during which many young people move out of the family home, begin establishing educational qualifications or transition to the labour market, and form important social relationships. We evaluate whether the impacts are modified by age or sex, mapping out the social risk factors for depression-overweight comorbidity. We use rich data from a UK birth cohort, including a validated depressive symptom questionnaire and measured BMI, and prospectively collected measures of socioeconomic indicators.

## METHODS

### Data

We use data from the Avon Longitudinal Study of Parents and Children (ALSPAC), a population-based prospective birth cohort (29–31). ALSPAC recruited 14,541 pregnant women residing in the former county of Avon around the city of Bristol in the South West of England, UK, with an estimated delivery date between April 1991 and December 1992. Of these initial pregnancies, 13,988 children were alive at 1 year of age. When the oldest children were approximately 7 years of age, an attempt was made to bolster the initial sample with eligible cases who had failed to join the study originally. The total sample size for analyses using any data collected after the age of seven is therefore 15,447 pregnancies, of which 14,901 children were alive at 1 year of age. The families have been followed up with regular assessments to the present day. The study website contains details of all the data that is available through a fully searchable data dictionary and variable search tool (http://www.bristol.ac.uk/alspac/researchers/our-data/).

In 2008-2011, at a target age of 17.5 years, 5,217 ALSPAC participants took part in a clinic where weight and height were measured, and 4,500 participants responded to an online questionnaire with data on depressive symptoms. A clinic in 2015-2017 was attended by 4,026 participants at approximately 24 years of age, and 4,222 ALSPAC participants completed a questionnaire including questions on depressive symptoms at approximately 23 years of age.

The sample for the present study includes participants who had data on depressive symptoms and/or BMI at the age 17 wave (Figure 1 in Supplemental Material). Study data were collected and managed using REDCap electronic data capture tools hosted at the University of Bristol.(32) REDCap (Research Electronic Data Capture) is a secure, web-based software platform designed to support data capture for research studies. Ethical approval for the study was obtained from the ALSPAC Ethics and Law Committee and the Local Research Ethics Committees. Informed consent for the use of data was obtained from participants following the recommendations of the ALSPAC Ethics and Law Committee at the time.

**Figure 1.**
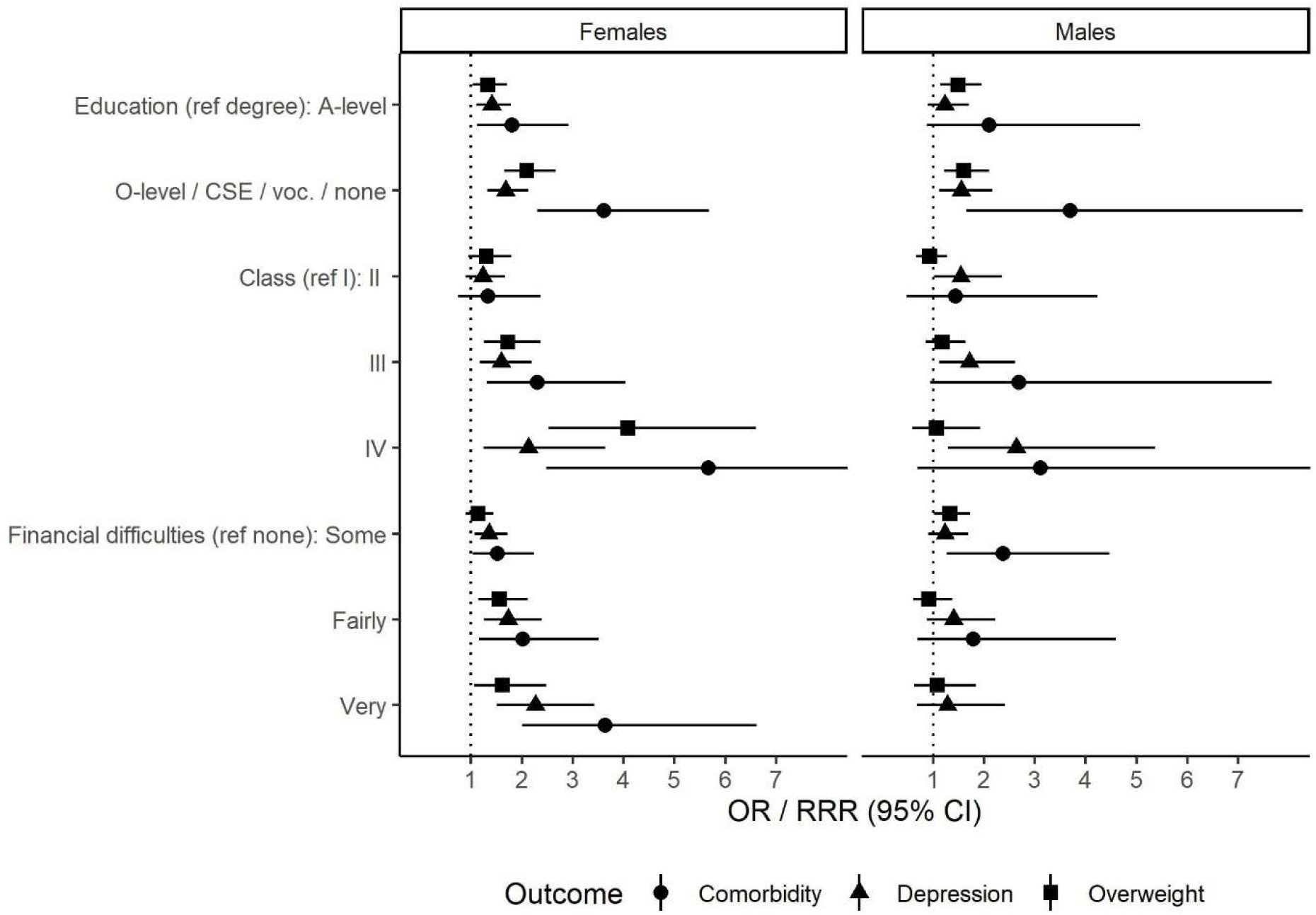
Association between family socioeconomic circumstances and depression, overweight and comorbidity at age 17 by sex. Note: Odds ratios (OR) from separate logistic regression models of depression versus no depression and overweight versus no overweight with each measure of socioeconomic circumstances, and relative risk ratios (RRR) from multinomial logistic regression models of comorbidity versus neither depression or overweight. Models adjusted for ethnicity. Model for comorbidity and financial difficulties in males did not converge due to low numbers.

### Outcomes

As our primary outcomes of interest at ages 17 and 24, we created multinomial outcome variables of comorbidity categorised as: (i) no depression or overweight (reference), (ii) depression only, (iii) overweight only, and (iv) depression and overweight comorbidity.

BMI (kg/m^2^) was calculated from measured weight and height. For ages under 18, BMI categories of ‘underweight ‘, ‘normal weight ‘, ‘overweight’ and ‘obesity’ were identified using sex- and age-specific BMI Z-scores relative to the UK1990 reference population (33), and at ages 18 and over, by BMI<18.5, BMI≥18.5 and <25, BMI≥25 and <30, and BMI≥30 respectively.

To identify depression, we used scores from the short Moods and Feelings Questionnaire (sMFQ), a 13-item self-reported questionnaire used for screening depressive symptoms in adolescents (34), administered through online or paper questionnaires. Participants were asked to rate statements about experiences of low mood and other correlates in the past two weeks as “not true”, “sometimes” or “true”. Total scores range between 0 and 26, with higher scores indicating more depressive symptoms. A score threshold of ≥ 11 was used to indicate depression (35,36).

### Exposures

Early-life socioeconomic circumstances Socioeconomic circumstances were measured with three indicators: parental education, social class and financial difficulties. Parental education was identified as the highest educational level from the mother ‘s and her partner ‘s self-reported qualifications in questionnaires during pregnancy, and categorised as (i) degree-level qualifications, (ii) Advanced (A)-level (examinations around age 18), and (iii) Ordinary (O)-level (examinations taking place at approx. age 16, the UK minimum school leaving age when the mothers were at school), CSE (Certificate of Secondary Education), vocational degree or no qualifications. Parental social class was derived from questionnaires during pregnancy and measured by the highest occupational class based on Registrar General ‘s Social Class classification: (i) I – professional, (ii) II – managerial/technical, and (ii) III – skilled manual or nonmanual, and (iv) IV – semiskilled manual or V – unskilled manual. Financial difficulties in affording food, heating, or rent or mortgage were assessed in a questionnaire at 32 weeks’ gestation. For each, mothers were asked “How difficult at the moment do you find it to afford these items” with the options “Very” “Fairly” “Slightly” or “Not difficult.” We constructed a multinomial categorical measure identifying if there were: (i) no difficulties reported, (ii) any reported as “slightly difficult” but not “fairly” or “very”, (iii) any reported as “fairly difficult” but not “very” and (iv) if any were reported as “very difficult.”

### Covariates

We included ethnicity reported by the mother (white/black or minority ethnicity) as a covariate. In additional models, we adjusted for maternal depressive score and BMI in early pregnancy as potential confounders, although they may mediate some of the effects of family SEP, as they are likely to be on the causal pathway between family SEP and adolescent health. Maternal depressive score was derived from a version of the Edinburgh Postnatal Depression Scale (EPDS) questionnaire at 32 weeks gestation (median 6.0, interquartile range 3.0-10.0). BMI was derived from self-reported pre-pregnancy weight and height in a questionnaire during or right after pregnancy (median BMI 22.1, interquartile range 20.5-24.2).

### Dealing with missing data

Of the participants who had at least one outcome measure at age 17 (N=4,948), 66% had any missing data in one or more of the exposures, outcomes or covariates. This varied from less than 3% for education or class, 6% for financial difficulties and 45% for depression at age 24 (Appendix Table 1). We used multiple imputation (MI) to increase power and reduce selection bias under a missing-at-random assumption. For imputation models, we included depression and BMI, all exposures and covariates from the analytical models, and additional variables that may be predictive of missingness or predict missing values themselves (Table 1 in Supplemental Material). We imputed data for males and females separately to enable comparisons across sexes. Comorbidity was passively imputed. We generated 50 imputed datasets with 20 cycles and combined coefficients across datasets using Rubin ‘s rules. MI was performed using the ‘mi impute chained’ command in Stata 17.

**Table 1.** Characteristics of the unimputed and imputed sample (N=4,948)

|  | **Unimputed** | **Imputed** |  |  |
| --- | --- | --- | --- | --- |
|  | **All** | **All** | **Females** | **Males** |
| **Sex, % (N)** |  |  |  |  |
| Male | 43.5 (2150) | 43.5 |  |  |
| Female | 56.5 (2798) | 56.5 |  |  |
| **Ethnicity, % (N)** |  |  |  |  |
| White | 95.7 (4557) | 95.6 | 95.5 | 95.6 |
| Other | 4.3 (206) | 4.4 | 4.4 | 4.4 |
| **Depression at 17, % (N)** |  |  |  |  |
| No | 78.5 (3332) | 78.2 | 74.2 | 83.5 |
| Yes | 21.5 (913) | 21.8 | 25.8 | 16.5 |
| **BMI category at 17, % (N)** |  |  |  |  |
| Underweight | 7.9 (373) | 7.9 | 7.8 | 8 |
| Normal weight | 69.3 (3279) | 69.2 | 68.2 | 70.4 |
| Overweight | 16.2 (766) | 16.3 | 16.7 | 15.8 |
| Obese | 6.6 (313) | 6.6 | 7.3 | 5.7 |
| **Comorbidity at 17, % (N)** |  |  |  |  |
| Has neither depression nor overweight | 61.9 (2493) | 62.1 | 59.2 | 65.9 |
| Depression only | 15.9 (640) | 15.6 | 17.7 | 13 |
| Overweight only | 16.9 (682) | 16.9 | 15.9 | 18.2 |
| Comorbidity | 5.3 (213) | 5.3 | 7.2 | 2.9 |
| **Depression at 23, % (N)** |  |  |  |  |
| No | 76.8 (2096) | 76.5 | 73.4 | 80.5 |
| Yes | 23.2 (632) | 23.5 | 26.6 | 19.5 |
| **BMI category at 24, % (N)** |  |  |  |  |
| Underweight | 3.0 (89) | 3.9 | 4.4 | 3.2 |
| Depression only | 59.6 (1754) | 55.8 | 55.9 | 55.5 |
| Overweight only | 24.5 (721) | 27.1 | 24.4 | 30.5 |
| Obese | 6.6 (313) | 13.3 | 15.3 | 10.8 |
| **Comorbidity at 24, % (N)** |  |  |  |  |
| Has neither depression nor overweight | 50.6 (1108) | 46.7 | 46.4 | 47.4 |
| Depression only | 12.9 (282) | 12.8 | 13.9 | 11.3 |
| Overweight only | 26.9 (588) | 30 | 27.1 | 33.1 |
| Comorbidity | 9.6 (211) | 10.7 | 12.6 | 8.2 |
| **Highest parental education, % (N)** |  |  |  |  |
| Degree | 29.3 (1423) | 29.2 | 27.8 | 31.1 |
| A-level | 35.5 (1721) | 35.4 | 35.2 | 35.6 |
| O-level / CSE / vocational / none | 35.2 (1707) | 35.4 | 37 | 33.3 |
| **Highest parental social class, % (N)** |  |  |  |  |
| I | 13.6 (653) | 13.5 | 12.6 | 14.7 |
| II | 45.0 (2166) | 44.9 | 44.6 | 45.2 |
| III | 37.4 (1802) | 37.6 | 38.8 | 36 |
| IV/V | 4.0 (191) | 4.1 | 4.1 | 4.1 |
| **Financial difficulties, % (N)** |  |  |  |  |
| No difficulties | 66.2 (3079) | 65.8 | 65.1 | 66.7 |
| Some difficulties | 20.8 (967) | 20.9 | 21.3 | 20.3 |
| Fairly difficult | 8.6 (401) | 8.8 | 9 | 8.4 |
| Very difficult | 4.4 (204) | 4.5 | 4.6 | 4.4 |
| **Maternal EPDS score, mean (SD)** | 6.55 (4.84) | 6.58 | 6.63 | 6.52 |
| **Maternal BMI, mean (SD)** | 22.84 (3.70) | 22.87 | 22.82 | 22.92 |

### Statistical methods

Analyses were conducted with Stata 17.0. First, we examined the sex-specific distributions, and the association between BMI category and depression with logistic regression models separately in girls and boys. We estimated odds ratios (ORs) for the associations between socioeconomic circumstances and depression or overweight with logistic regression. In the main analysis, we modelled depression-overweight comorbidity using multinomial logistic regression models which return relative risk ratios (RRRs). We adjusted for ethnicity and ran the models separately by sex and age and included each exposure (parental education, social class and financial difficulties) separately. We tested the interactions between socioeconomic measures and sex with joint tests of interaction terms, applying a threshold of p<0.05 for statistical evidence of interaction. We also used the user-written Stata command ‘riigen’ to construct relative indices of inequality (RIIs) for each socioeconomic measure to enable more direct comparison. This converts the exposures into continuous variables representing the proportion of the sample with greater socioeconomic disadvantage than that group to compare the top and bottom of a hypothetical distribution of a continuous SEP marker. In sensitivity analyses, we adjusted for maternal depressive score and BMI. We also repeated the main analyses (a) using BMI≥25 as the definition of overweight at age 17 for all, (b) using depression-obesity comorbidity as the outcome, and (c) in the unimputed data.

## RESULTS

The characteristics of the unimputed and imputed data were highly similar (Table 1), and we report results using imputed data. Females were more likely to have depression than males regardless of having overweight. Overall, 25.8% of females and 16.5% of males overall had depression at age 17, and of those with overweight, 31.3% had depression in females, and 13.7% in males. Proportions with overweight or obesity at age 17 were similar by sex, while 7.2% of girls and 2.9% of boys had depression and overweight comorbidity. The prevalence of overweight and obesity increased substantially from age 17 to 24, and depression and comorbidity prevalence increased as well. At age 24, 12.6% of females and 8.2% of males had comorbidity. Overweight and particularly obesity was associated with greater odds of depression in females, while in males the estimates were imprecise with no clear relationship (Table 2).

**Table 2.** Association between BMI category and depression at ages 17 and 24.

|  | **Females** |  | **Males** |  |
| --- | --- | --- | --- | --- |
| **Age 17** | **OR** | **95% CI** | **OR** | **95% CI** |
| Underweight | 1.39 | 1.00, 1.92 | 1.17 | 0.76, 1.82 |
| Normal weight | ref |  | ref |  |
| Overweight | 1.31 | 1.02, 1.69 | 0.79 | 0.54, 1.16 |
| Obesity | 1.95 | 1.38, 2.75 | 0.79 | 0.43, 1.43 |
| **Age 24** |  |  |  |  |
| Underweight | 1.01 | 0.60, 1.70 | 0.94 | 0.38, 2.35 |
| Normal weight | ref |  | ref |  |
| Overweight | 1.37 | 1.06, 1.78 | 0.97 | 0.67, 1.40 |
| Obesity | 1.88 | 1.38, 2.57 | 1.24 | 0.74, 2.08 |

Note: Models adjusted for ethnicity.

Figure 1 demonstrates associations of the three measures of socioeconomic circumstances with depression, overweight and comorbidity at age 17. Low parental education, class and greater financial difficulties were all associated with greater odds of depression in females (Figure 1, Supplemental Material Table 2). In males, results for education and class were similar to those seen with girls, but results with financial difficulties were weaker. All three measures were associated with the odds of overweight at age 17 in females (Figure 1, Supplemental Material Table 3), and particularly the lowest parental social class (IV/V) demonstrated a strong association with having overweight. There was less evidence of an association between class or financial difficulties with overweight in males.

Estimates of the associations with comorbidity were greater than those for depression or overweight only (Table 4 in Supplemental Material). Low education and social class were associated with greater RRR of comorbidity in both sexes (Figure 1). Financial difficulties were a risk factor for comorbidity in females, but the pattern was less clear in males. Testing the evidence for differential SEP effects by sex, we found no statistically significant differences but interaction terms were imprecise (Table 5 in Supplemental Material), but we report the results by sex due to the differential association of depression and overweight by sex. Using RII to compare, low education was the strongest risk factor for comorbidity of the three measures in both girls and boys (Table 6 in Supplemental Material).

At age 24 (Figure 2, Supplemental Material Table 2, 3, 4), the results for depression, overweight and comorbidity were very similar. Education remained the strongest risk factor for comorbidity for females at age 24, but the RII for class was slightly higher for males (Table 6 in Supplemental Material).

**Figure 2.**
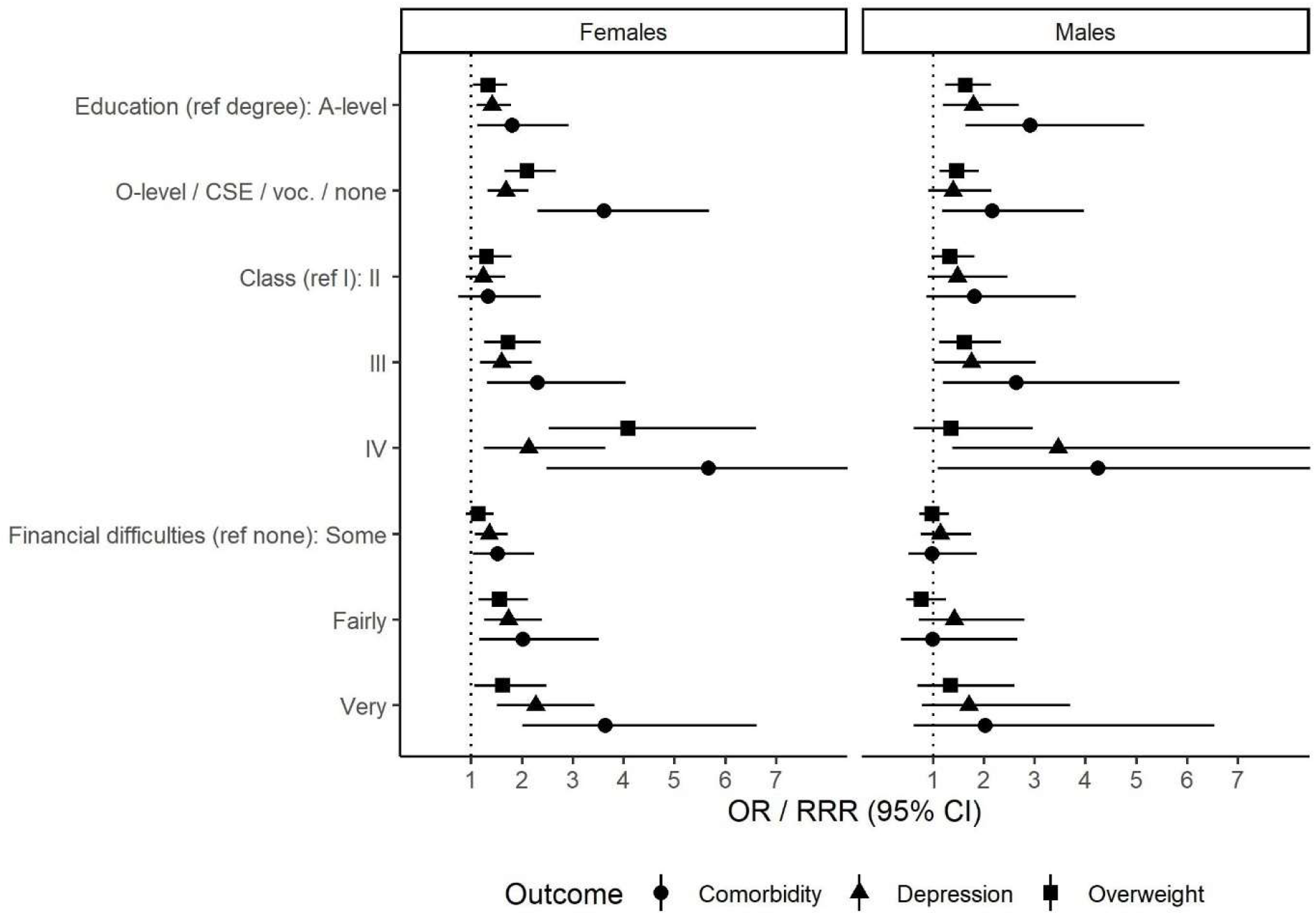
Association between family socioeconomic circumstances and depression, overweight and comorbidity at age 24 by sex. Note: Odds ratios (OR) from separate logistic regression models of depression versus no depression and overweight versus no overweight with each measure of socioeconomic circumstances, and relative risk ratios (RRR) from multinomial logistic regression models of comorbidity versus neither depression or overweight. Models adjusted for ethnicity.

### Sensitivity analyses

Results additionally adjusting for maternal depressive symptoms and BMI attenuated the OR for comorbidity by between 22 to 42% for education, 27 to 55% for class, and 13 to 48% for financial difficulties at age 17, but did not remove them (Supplemental Material Table 7). Using BMI≥25 definition of overweight at age 17 showed similar results (Supplemental Material Table 8). Defining comorbidity with obesity rather than overweight generally showed slightly greater risk associated with socioeconomic measures, but confidence intervals were wider (Supplemental Material Table 9). Finally, results for the main models in unimputed data were highly similar to the imputed data (Supplemental Material Table 10).

## DISCUSSION

Using data of adolescents coming of age in UK in the 2000s, we found that early-life parental socioeconomic circumstances were associated with depression, overweight and their comorbidity in adolescence and young adulthood. Parental education, social class and financial difficulties were all risk factors for comorbidity, and generally more strongly for comorbidity than depression or overweight only. The findings were similar from adolescence to young adulthood, and we did not find strong evidence of sex differences.

The comorbidity of obesity and depression is likely to be one of the earliest life course manifestations of multimorbidity, with potentially serious lifelong implications for subsequent chronic diseases and wellbeing. The findings contribute to our understanding of the early social causes by comparing relationships of different socioeconomic risk factors. Our results corroborate previous findings of an association between parental social class and comorbidity (13). However, our results indicate that parental education tended to be the strongest indicator of comorbidity risk, demonstrated by the higher RII values. Parental education influences several aspects of socioeconomic circumstances such as occupation, income, financial difficulties, housing and neighbourhood, but may also be more strongly associated with physical activity and diet, which are likely to have an impact on depression-overweight comorbidity. Mirowsky and Ross suggested that self-efficacy plays a part in the effects of education on health (37). More educated parents may have the resources to apply a greater emphasis on fostering health promoting behaviours in the family environment.

The relationships between social class or financial difficulties and the outcome indicate the impact of early-life material conditions. We did not have a measure of family income, but self-reported financial difficulties in affording food, heating and accommodation capture significant aspects of poverty. Experiencing poverty can be a substantial source of stress that may influence parenting and contribute to household conflict (38), and reduce emotional coping resources as well as material resources for healthy life styles. We used a measure of financial difficulties from early-life, but it is possible that financial difficulties experienced in adolescence would have a greater impact on concurrent mental and physical health problems and should be explored further. Irrespective, an increase in families with children experiencing financial difficulties is likely to have significant short and long term effects on adolescent health and wellbeing.

We presented the results by sex, because females show a stronger association between depression and overweight and greater prevalence of comorbidity. These results indicate patterns and processes influenced by sex and/or gender, such as experiences of weight stigma and body satisfaction (28). However, we did not find statistical evidence for sex differences in the impact of SEP, which is in agreement with a previous study using earlier British cohorts examining father ‘s social class and comorbidity in childhood (13).

An advantage of the study was its longitudinal study design, with socioeconomic measures reported in early-life by mothers and the follow up through adolescence and young adulthood. BMI was measured by trained staff, and the measure of depression used is likely to be more sensitive and less influenced by socioeconomic background than relying on a clinical diagnosis of depression from primary care data which may be influenced by the likelihood of seeking care. However, one of the limitations of the study was the potential bias and low precision in the results due to study attrition and missingness on individual items. Loss to follow-up in ALSPAC is not random (39) and those with poorer health and lower SEP are more likely to drop out, which can bias estimates of SEP differences in health towards the null (40). We mitigated this problem by using the maximum sample available, i.e. those with any outcome data at age 17, and applying MI for missing data. We found that results in the unimputed and imputed data were similar. Nevertheless, ALSPAC remains a more advantaged sample than the general English population at the time and predominantly white (30), which may have an impact on generalizability of the findings.

In conclusion, socioeconomic circumstances in early-life predict depression and overweight comorbidity in adolescence and young adulthood, highlighting the early emergence of socioeconomic inequalities in multimorbidity. Future research should ascertain how family SEP is associated with the longitudinal reciprocal relationship between BMI and depressive symptoms in adolescence.

## Data Availability

The informed consent obtained from ALSPAC participants does not allow the data to be made freely available through any third party maintained public repository. However, data used for this submission can be made available on request to the ALSPAC Executive. The ALSPAC data management plan describes in detail the policy regarding data sharing, which is through a system of managed open access. Full instructions for applying for data access can be found here: http://www.bristol.ac.uk/alspac/researchers/access/. The ALSPAC study website contains details of all the data that are available (http://www.bristol.ac.uk/alspac/researchers/our-data/).

## Acknowledgements

We are extremely grateful to all the families who took part in this study, the midwives for their help in recruiting them, and the whole ALSPAC team, which includes interviewers, computer and laboratory technicians, clerical workers, research scientists, volunteers, managers, receptionists and nurses.

## Funding

FK was funded by an Economic and Social Research Council grant (ES/T013923/1). The UK Medical Research Council and Wellcome (Grant ref: 217065/Z/19/Z) and the University of Bristol provide core support for ALSPAC. A comprehensive list of grants funding is available on the ALSPAC website (http://www.bristol.ac.uk/alspac/external/documents/grant-acknowledgements.pdf); The collection of some of the variables used in this study were specifically funded by Wellcome Trust and MRC (076467/Z/05/Z and MR/L022206/1). This publication is the work of the authors and FK will serve as guarantor for the contents of this paper.

## Competing interests

FK declares no competing interests. LDH declares grants from the Medical Research Council and Health Foundation and membership of Academy of Finland funding panel.

## Notes

### Author Declarations

Ethical approval for the study was obtained from the ALSPAC Ethics and Law Committee and the Local Research Ethics Committees. Informed consent for the use of data was obtained from participants following the recommendations of the ALSPAC Ethics and Law Committee at the time.

## REFERENCES

1. Fergusson DM, Boden JM, Horwood LJ. Recurrence of major depression in adolescence and early adulthood, and later mental health, educational and economic outcomes. Br J Psychiatry. 2007 Oct 2;191(04):335–42.

2. Singh AS, Mulder C, Twisk JWR, Van Mechelen W, Chinapaw MJM. Tracking of childhood overweight into adulthood: a systematic review of the literature. Obes Rev. 2008 Mar 5;9(5):474–88.

3. Norris T, Bann D, Hardy R, Johnson W. Socioeconomic inequalities in childhood-to-adulthood BMI tracking in three British birth cohorts. Int J Obes 2019 442. 2019 Jun 5;44(2):388–98.

4. Milaneschi Y, Simmons WK, van Rossum EFC, Penninx BW. Depression and obesity: evidence of shared biological mechanisms. Mol Psychiatry. 2019 Jan;24(1):18–33.

5. Konttinen H, Kiviruusu O, Huurre T, Haukkala A, Aro H, Marttunen M. Longitudinal associations between depressive symptoms and body mass index in a 20-year followup. Int J Obes. 2014 May 16;38(5):668–74.

6. Davillas A, Benzeval M, Kumari M. Association of Adiposity and Mental Health Functioning across the Lifespan: Findings from Understanding Society (The UK Household Longitudinal Study). Meyre D, editor. PLoS One. 2016 Feb 5;11(2):e0148561.

7. Sutaria S, Devakumar D, Yasuda SS, Das S, Saxena S. Is obesity associated with depression in children? Systematic review and meta-analysis. Arch Dis Child. 2018 Jun 29;104(1):archdischild-2017–314608.

8. Luppino FS, De Wit LM, Bouvy PF, Stijnen T, Cuijpers P, Penninx BWJH, et al. Overweight, obesity, and depression: A systematic review and meta-analysis of longitudinal studies. Vol. 67, Archives of General Psychiatry. American Medical Association; 2010. p. 220–9.

9. de Wit L, Luppino F, van Straten A, Penninx B, Zitman F, Cuijpers P. Depression and obesity: A meta-analysis of community-based studies. Psychiatry Res. 2010 Jul 30;178(2):230–5.

10. Stunkard AJ, Faith MS, Allison KC. Depression and obesity. Biol Psychiatry. 2003 Aug 1;54(3):330–7.

11. Mannan M, Mamun A, Doi S, Clavarino A. Prospective Associations between Depression and Obesity for Adolescent Males and Females-A Systematic Review and Meta-Analysis of Longitudinal Studies. Homberg J, editor. PLoS One. 2016 Jun 10;11(6):e0157240.

12. Patalay P, Hardman CA. Comorbidity, Codevelopment, and Temporal Associations Between Body Mass Index and Internalizing Symptoms From Early Childhood to Adolescence. JAMA Psychiatry. 2019 Mar 20;

13. Khanolkar AR, Patalay P. Socioeconomic inequalities in co-morbidity of overweight, obesity and mental ill-health from adolescence to mid-adulthood in two national birth cohort studies. Lancet Reg Heal - Eur. 2021;6.

14. Joinson C, Kounali D, Lewis G. Family socioeconomic position in early life and onset of depressive symptoms and depression: a prospective cohort study. Soc Psychiatry Psychiatr Epidemiol. 2017 Jan 11;52(1):95–103.

15. Björkenstam E, Pebley AR, Burström B, Kosidou K. Childhood social adversity and risk of depressive symptoms in adolescence in a US national sample. J Affect Disord. 2017 Apr 23;212:56–63.

16. Wirback T, Möller J, Larsson JO, Galanti MR, Engström K. Social factors in childhood and risk of depressive symptoms among adolescents - a longitudinal study in Stockholm, Sweden. Int J Equity Health. 2014;13(1):1–11.

17. Reiss F. Socioeconomic inequalities and mental health problems in children and adolescents: A systematic review. Vol. 90, Social Science and Medicine. Soc Sci Med; 2013. p. 24–31.

18. Wardle J, Brodersen NH, Cole TJ, Jarvis MJ, Boniface DR. Development of adiposity in adolescence: five year longitudinal study of an ethnically and socioeconomically diverse sample of young people in Britain. BMJ. 2006 May 13;332(7550):1130–5.

19. Shrewsbury V, Wardle J. Socioeconomic Status and Adiposity in Childhood: A Systematic Review of Cross-sectional Studies 1990–2005. Obesity. 2008 Feb 1;16(2):275–84.

20. Barriuso L, Miqueleiz E, Albaladejo R, Villanueva R, Santos JM, Regidor E. Socioeconomic position and childhoodadolescent weight status in rich countries: A systematic review, 1990–2013. BMC Pediatr. 2015 Sep 21;15(1):1–15.

21. Link BG, Phelan J. Social Conditions As Fundamental Causes of Disease. J Health Soc Behav. 1995;35:80.

22. Braveman PA, Cubbin C, Egerter S, Chideya S, Marchi KS, Metzler M, et al. Socioeconomic Status in Health Research. JAMA. 2005 Dec 14;294(22):2879.

23. Geyer S, Hemström O, Peter R,Vågerö D. Education, income, and occupational class cannot be used interchangeably in social epidemiology. Empirical evidence against a common practice. J Epidemiol Community Heal. 2006;60(9):804–10.

24. Sobal J. Obesity and socioeconomic status: A framework for examining relationships between physical and social variables. Med Anthropol. 1991;13:231–47.

25. Pampel FC, Krueger PM, Denney JT. Socioeconomic Disparities in Health Behaviors. Annu Rev Sociol. 2010 Jun;36(1):349–70.

26. Kuehner C. Why is depression more common among women than among men? Vol. 4, The Lancet Psychiatry. Elsevier Ltd; 2017. p. 146–58.

27. Tseng LA, El Khoudary SR, Young EA, Farhat GN, Sowers M, Sutton-Tyrrell K, et al. The association of menopause status with physical function. Menopause J North Am Menopause Soc. 2012 Nov;19(11):1186–92.

28. Puhl RM, Heuer CA. The Stigma of Obesity: A Review and Update. Obesity. 2009 May;17(5):941–64.

29. Boyd A, Golding J, Macleod J, Lawlor DA, Fraser A, Henderson J, et al. Cohort Profile: The ‘Children of the 90s ‘—the index offspring of the Avon Longitudinal Study of Parents and Children. Int J Epidemiol. 2013 Feb 1;42(1):111–27.

30. Fraser A, Macdonald-Wallis C, Tilling K, Boyd A, Golding J, Davey Smith G, et al. Cohort Profile: The Avon Longitudinal Study of Parents and Children: ALSPAC mothers cohort. Int J Epidemiol. 2013 Feb 1;42(1):97–110.

31. Northstone K, Lewcock M, Groom A, Boyd A, Macleod J, Timpson N, et al. The Avon Longitudinal Study of Parents and Children (ALSPAC): an update on the enrolled sample of index children in 2019. Wellcome open Res. 2019;4:51.

32. Harris PA, Taylor R, Thielke R, Payne J, Gonzalez N, Conde JG. Research electronic data capture (REDCap)-A metadata-driven methodology and workflow process for providing translational research informatics support. J Biomed Inform. 2009 Apr;42(2):377–81.

33. Vidmar SI, Cole TJ, Pan H. Standardizing anthropometric measures in children and adolescents with functions for egen: Update. Stata J. 2013 Jul 1;13(2):366–78.

34. Angold A, Costello J, Van Kämmen W, Stouthamer-Loeber M. Development of a short questionnaire for use in epidemiological studies of depression in children and adolescents: factor composition and structure across development. Int J Methods Psychiatr Res. 1996;5(4):251–62.

35. Kwong ASF. Examining the longitudinal nature of depressive symptoms in the Avon Longitudinal Study of Parents and Children (ALSPAC). Wellcome Open Res. 2019 Oct 4;4:126.

36. Turner N, Joinson C, Peters TJ, Wiles N, Lewis G. Validity of the Short Mood and feelings questionnaire in late adolescence. Psychol Assess. 2014;26(3):752–62.

37. Mirowsky J, Ross CE. Education, Social Status and Health. New York: Aldine de Gruyter; 2003.

38. Conger RD, Conger KJ, Elder Jr. GH Lorenz FO, Simons RL, Whitbeck LB. A Family Process Model of Economic Hardship and Adjustment of Early Adolescent Boys. Child Dev. 1992 Jun 1;63(3):526–41.

39. Cornish RP, MacLeod J, Boyd A, Tilling K. Factors associated with participation over time in the Avon Longitudinal Study of Parents and Children: a study using linked education and primary care data. Int J Epidemiol. 2021 Mar 3;50(1):293–302.

40. Howe LD, Tilling K, Galobardes B, Lawlor DA. Loss to Follow-up in Cohort Studies: Bias in Estimates of Socioeconomic Inequalities. Epidemiology. 2013 Jan;24(1):1.

