## Supplemental Material for "Early-life socioeconomic circumstances and the comorbidity of depression and overweight in adolescence and young adulthood: a longitudinal study"

Contents:

Figure 1. Flow diagram of sample selection

Table 1. Missing data and model for multiple imputation

Table 2. Association between family socioeconomic circumstances and depression at ages 17 and 24

Table 3. Association between family socioeconomic circumstances and overweight at ages 17 and 24

Table 4. Association between family socioeconomic circumstances and overweight-depression comorbidity versus neither depression or overweight at ages 17 and 24

Table 5. Interaction between sex and socioeconomic circumstances and overweight-depression comorbidity versus neither depression or overweight at ages 17 and 24

Table 6. Relative indices of inequality for education, class and financial difficulties

Table 7. Association between family socioeconomic circumstances and comorbidity at ages 17 and 24 adjusted for additionally for maternal BMI and depressive score

Table 8. Association between family socioeconomic circumstances and depression-overweight comorbidity defined by BMI>25 at ages 17 and 24

Table 9. Association between family socioeconomic circumstances and depression and obesity comorbidity at ages 17 and 24

Table 10. Association between family socioeconomic circumstances and depression-overweight comorbidity at ages 17 and 24 in unimputed sample

**Figure 1. Flow diagram of sample selection**

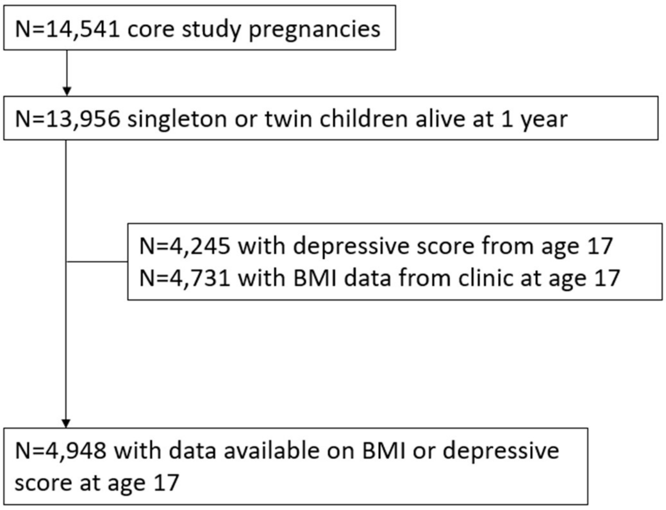

**Table 1. Missing data and model for multiple imputation**

| Variable | Type | Missing data (%) | Model | Imputation model covariates |
|---|---|---|---|---|
| Sex | covariate | 0 | NA | NA |
| Ethnicity | covariate | 4 | logistic | education, class, financial difficulties, logged BMI at 17, depression at 17, logged BMI at 24, depression at 23, maternal depressive score, maternal BMI, birthweight, gestational age, age of mother, parity, mother's smoking in pregnancy, housing tenure, parents' marital status |
| Parental education | exposure | 2 | multinomial logistic | social class, financial difficulties, logged BMI at 17, depression at 17, logged BMI at 24, depression at 23, ethnicity, maternal depressive score, maternal BMI, birthweight, gestational age, age of mother, parity, mother's smoking in pregnancy, housing tenure, parents' marital status |
| Social class | exposure | 3 | multinomial logistic | education, financial difficulties, logged BMI at 17, depression at 17, logged BMI at 24, depression at 23, ethnicity, maternal depressive score, maternal BMI, birthweight, gestational age, age of mother, parity, mother's smoking in pregnancy, housing tenure, parents' marital status |
| Financial difficulties | exposure | 6 | multinomial logistic | education, social class, logged BMI at 17, depression at 17, logged BMI at 24, depression at 23, ethnicity, maternal depressive score, maternal BMI, birthweight, gestational age, age of mother, parity, mother's smoking in pregnancy, housing tenure, parents' marital status |
| Logged BMI at age 17 | outcome | 4 | linear | overweight at age 15, depression at 17, depression at 23, education, class, financial difficulties, ethnicity, maternal depressive score, maternal BMI, birthweight, gestational age, age of mother, parity, mother's smoking in pregnancy, housing tenure, parents' marital status |
| Depression at age 17 | outcome | 14 | logistic | depression at ages 16, 18, 21 and 22, logged BMI at 17, logged BMI at 24, education, class, financial difficulties, ethnicity, maternal depressive score, maternal BMI, , birthweight, gestational age, age of mother, parity, mother's smoking in pregnancy, housing tenure, parents' marital status |
| Logged BMI at age 24 | outcome | 41 | linear | overweight at age 15, depression at 17, depression at 23, education, class, financial difficulties, ethnicity, maternal depressive score, maternal BMI, birthweight, gestational age, age of mother, parity, mother's smoking in pregnancy, housing tenure, parents' marital status |
| Depression at age 23 | outcome | 45 | logistic | depression at ages 16, 18, 21 and 22, logged BMI at 17, logged BMI at 24, education, class, financial difficulties, ethnicity, maternal depressive score, maternal BMI, , birthweight, gestational age, age of mother, parity, mother's smoking in pregnancy, housing tenure, parents' marital status |
| Maternal BMI | covariate | 9 | linear | education, class, financial difficulties, ethnicity, logged BMI at 17, depression at 17, logged BMI at 24, depression at 23, maternal depressive score, birthweight, gestational age, age of mother, parity, mother's smoking in pregnancy, housing tenure, parents' marital status |
| Maternal depressive score | covariate | 6 | linear | education, class, financial difficulties, ethnicity, logged BMI at 17, depression at 17, logged BMI at 24, depression at 23, maternal BMI, birthweight, gestational age, age of mother, parity, mother's smoking in pregnancy, housing tenure, parents' marital status |

**Table 2. Association between family socioeconomic circumstances and depression at ages 17 and 24**

| | **Female age 17** | | **Female age 24** | | **Male age 17** | | **Male age 24** | |
|---|---|---|---|---|---|---|---|---|
| **EDUCATION** | **OR** | **95% CI** | **OR** | **95% CI** | **OR** | **95% CI** | **OR** | **95% CI** |
| | ref | | ref | | ref | | ref | |
| Degree | 1.4 | 1.10, 1.78 | 1.57 | 1,20, 2.04 | 1.23 | 0.89,1.70 | 1.79 | 1.19, 2.69 |
| A-level | 1.68 | 1.32, 2.13 | 1.29 | 0.98, 1.69 | 1.56 | 1.12, 2.16 | 1.39 | 0.90, 2.15 |
| O-level / CSE / vocational / none | | | | | | | | |
| **CLASS** | ref | | ref | | ref | | ref | |
| I | 1.23 | 0.90, 1.68 | 0.99 | 0.71, 1.40 | 1.55 | 1.02, 2.35 | 1.48 | 0.89, 2.46 |
| II | 1.6 | 1.17, 2.19 | 1.27 | 0.90, 1.81 | 1.71 | 1.12, 2.62 | 1.75 | 1.01, 3.02 |
| III | 2.13 | 1.24, 3.64 | 1.19 | 0.56, 2.52 | 2.63 | 1.29, 5.36 | 3.45 | 1.37, 8.69 |
| IV/V | | | | | | | | |
| **FINANCIAL DIFFICULTIES** | | | | | | | | |
| None reported | ref | | ref | | ref | | ref | |
| Some | 1.35 | 1.07, 1.71 | 1.34 | 1.03, 1.75 | 1.24 | 0.90, 1.69 | 1.15 | 0.75, 1.75 |
| Fairly difficult | 1.74 | 1.26, 2.39 | 1.64 | 1.14, 2.35 | 1.4 | 0.88, 2.23 | 1.42 | 0.71, 2.80 |
| Very difficult | 2.27 | 1.50, 3.43 | 1.32 | 0.77, 2.25 | 1.29 | 0.68, 2.42 | 1.7 | 0.78, 3.70 |

Note: Logistic regression models adjusted for ethnicity.

**Table 3. Association between family socioeconomic circumstances and overweight at ages 17 and 24**

| | Female age 17 | | Female age 24 | | Male age 17 | | Male age 24 | |
|---|---|---|---|---|---|---|---|---|
| **EDUCATION** | **OR** | **95% CI** | **OR** | **95% CI** | **OR** | **95% CI** | **OR** | **95% CI** |
| Degree | ref | | ref | | ref | | | |
| A-level | 1.33 | 1.04, 1.71 | 1.62 | 1.28, 2.06 | 1.49 | 1.13, 1.95 | 1.63 | 1.25, 2.14 |
| O-level / CSE / vocational / none | 2.1 | 1.66, 2.66 | 2.32 | 1.84, 2.93 | 1.60 | 1.22, 2.10 | 1.47 | 1.13, 1.91 |
| **CLASS** | | | | | | | | |
| I | ref | | ref | | ref | | ref | |
| II | 1.3 | 0.95, 1.79 | 1.45 | 1.08, 1.94 | 0.93 | 0.67, 1.28 | 1.33 | 0.97, 1.82 |
| III | 1.73 | 1.26, 2.37 | 2.1 | 1.54, 2.87 | 1.18 | 0.85, 1.64 | 1.62 | 1.12, 2.33 |
| IV/V | 4.09 | 2.53, 6.60 | 3.65 | 2.05, 6.49 | 1.07 | 0.59, 1.93 | 1.35 | 0.62, 2.96 |
| **FINANCIAL DIFFICULTIES** | | | | | | | | |
| None reported | ref | | ref | | ref | | ref | |
| Some | 1.14 | 0.90, 1.44 | 0.97 | 0.77, 1.23 | 1.33 | 1.02, 1.73 | 0.98 | 0.73, 1.32 |
| Fairly difficult | 1.55 | 1.14, 2.11 | 1.45 | 1.04, 2.02 | 0.92 | 0.61, 1.38 | 0.77 | 0.47, 1.25 |
| Very difficult | 1.62 | 1.06, 2.47 | 2.04 | 1.31, 3.18 | 1.08 | 0.63, 1.84 | 1.34 | 0.69, 2.60 |

Note: Logistic regression models adjusted for ethnicity.

**Table 4. Association between family socioeconomic circumstances and overweight-depression comorbidity versus neither depression or overweight at ages 17 and 24**

| | Female age 17 | | Female age 24 | | Male age 17 | | Male age 24 | |
|---|---|---|---|---|---|---|---|---|
| **EDUCATION** | | | | | | | | |
| **Depression only** | RRR | 95% CI | RRR | 95% CI | RRR | 95% CI | RRR | 95% CI |
| Degree | ref | | ref | | ref | | ref | |
| A-level | 1.4 | 1.06,1.83 | 1.42 | 1.02,1.97 | 1.26 | 0.88,1.80 | 1.84 | 1.12,3.01 |
| O-level / CSE / vocational / none | 1.5 | 1.14,1.97 | 1.14 | 0.79,1.64 | 1.51 | 1.04,2.19 | 1.31 | 0.78,2.21 |
| **Overweight only** | | | | | | | | |
| Degree | ref | | ref | | ref | | ref | |
| A-level | 1.32 | 0.99,1.77 | 1.51 | 1.14,2.00 | 1.5 | 1.12,2.00 | 1.66 | 1.22,2.25 |
| O-level / CSE / vocational / none | 1.9 | 1.44,2.50 | 2.23 | 1.70,2.92 | 1.54 | 1.14,2.07 | 1.43 | 1.06,1.92 |
| **Both depression and overweight** | | | | | | | | |
| Degree | ref | | ref | | ref | | ref | |
| A-level | 1.81 | 1.12,2.91 | 2.5 | 1.69,3.69 | 2.11 | 0.88,5.06 | 2.91 | 1.64,5.15 |
| O-level / CSE / vocational / none | 3.61 | 2.30,5.67 | 2.86 | 1.93,4.23 | 3.69 | 1.65,8.26 | 2.17 | 1.19,3.97 |
| **CLASS** | | | | | | | | |
| **Depression only** | | | | | | | | |
| I | ref | | ref | | ref | | ref | |
| II | 1.3 | 0.90,1.87 | 0.95 | 0.63,1.41 | 1.61 | 1.01,2.56 | 1.59 | 0.84,3.01 |
| III | 1.61 | 1.11,2.33 | 1.21 | 0.80,1.83 | 1.63 | 1.01,2.63 | 1.84 | 0.93,3.63 |
| IV/V | 2.74 | 1.35,5.56 | 1.09 | 0.40,3.00 | 2.37 | 1.05,5.36 | 3.81 | 1.27,11.42 |
| **Overweight only** | | | | | | | | |
| I | ref | | ref | | ref | | ref | |
| II | 1.36 | 0.93,1.98 | 1.45 | 1.02,2.04 | 0.95 | 0.67,1.33 | 1.37 | 0.96,1.95 |
| III | 1.73 | 1.19,2.54 | 2.1 | 1.46,3.03 | 1.15 | 0.81,1.62 | 1.65 | 1.11,2.44 |
| IV/V | 5.13 | 2.82,9.34 | 3.7 | 1.83,7.48 | 1.06 | 0.54,2.07 | 1.42 | 0.56,3.59 |
| **Both depression and overweight** | | | | | | | | |
| I | ref | | ref | | ref | | ref | |
| II | 1.33 | 0.75,2.37 | 1.42 | 0.85,2.37 | 1.44 | 0.49,4.23 | 1.81 | 0.86,3.80 |
| III | 2.3 | 1.31,4.04 | 2.44 | 1.44,4.13 | 2.68 | 0.94,7.66 | 2.64 | 1.19,5.85 |
| IV/V | 5.67 | 2.48,12.94 | 3.77 | 1.46,9.74 | 3.11 | 0.70,13.91 | 4.24 | 1.10,16.40 |
| **FINANCIAL DIFFICULTIES** | | | | | | | | |
| **Depression only** | | | | | | | | |
| None reported | ref | | ref | | ref | | ref | |
| Some | 1.26 | 0.95,1.68 | 1.42 | 1.00,2.02 | 1.08 | 0.75,1.56 | 1.31 | 0.80,2.13 |
| Fairly difficult | 1.99 | 1.37,2.87 | 1.41 | 0.86,2.31 | 1.32 | 0.80,2.19 | 1.53 | 0.73,3.20 |
| Very difficult | 2.1 | 1.28,3.46 | 1.24 | 0.52,2.97 | 1.33 | 0.66,2.67 | 1.97 | 0.72,5.42 |
| **Overweight only** | | | | | | | | |
| None reported | ref | | ref | | ref | | ref | |

| | | | | | | | | |
|---|---|---|---|---|---|---|---|---|
| Some | 1.07 | 0.80,1.42 | 0.99 | 0.75,1.31 | 1.21 | 0.90,1.63 | 1.04 | 0.75,1.44 |
| Fairly difficult | 1.83 | 1.25,2.67 | 1.29 | 0.86,1.92 | 0.83 | 0.52,1.33 | 0.79 | 0.46,1.37 |
| Very difficult | 1.32 | 0.73,2.38 | 2.03 | 1.18,3.49 | 1.18 | 0.67,2.09 | 1.47 | 0.74,2.94 |
| **Both depression and overweight** | | | | | | | | |
| None reported | ref | | ref | | ref | | ref | |
| Some | 1.52 | 1.04,2.25 | 1.23 | 0.85,1.79 | 2.38 | 1.26,4.47 | 0.98 | 0.52,1.86 |
| Fairly difficult | 2.01 | 1.15,3.52 | 2.28 | 1.41,3.68 | 1.78 | 0.69,4.59 | 0.99 | 0.37,2.66 |
| Very difficult | 3.64 | 2.01,6.61 | 2.46 | 1.24,4.85 | 0 | - | 2.02 | 0.63,6.53 |

Note: Multinomial logistic regression models adjusted for ethnicity.

**Table 5. Interaction between sex and socioeconomic circumstances and overweight-depression comorbidity versus neither depression or overweight at ages 17 and 24**

| | | Age 17 | | | Age 24 | | |
|---|---|---|---|---|---|---|---|
| | | **RR** | **95% CI** | **P-value** | **RR** | **95% CI** | **P-value** |
| **Education** | | | | | | | |
| | **Depression only** | | | | | | |
| Main effects | Female | 1.45 | 1.05,2.02 | | 1.45 | 0.94,2.23 | |
| | A-level | 1.26 | 0.88,1.80 | | 1.84 | 1.13,3.01 | |
| | O-level/CSE/vocational/none | 1.51 | 1.04,2.19 | | 1.31 | 0.78,2.21 | |
| Interaction terms | Female X A-level | 1.11 | 0.71,1.74 | | 0.77 | 0.42,1.39 | |
| | Female X O-level/etc. | 0.99 | 0.62,1.58 | | 0.87 | 0.47,1.59 | |
| | **Overweight only** | | | | | | |
| Main effects | Female | 0.93 | 0.68,1.27 | | 0.71 | 0.53,0.95 | |
| | A-level | 1.5 | 1.12,2.00 | | 1.65 | 1.22,2.25 | |
| | O-level/CSE/vocational/none | 1.54 | 1.14,2.07 | | 1.42 | 1.06,1.91 | |
| Interaction terms | Female X A-level | 0.88 | 0.59,1.33 | | 0.91 | 0.60,1.37 | |
| | Female X O-level/etc. | 1.23 | 0.82,1.85 | | 1.57 | 1.06,2.33 | |
| | **Comorbidity** | | | | | | |
| Main effects | Female | 2.91 | 1.30,6.55 | | 1.49 | 0.85,2.59 | |
| | A-level | 2.1 | 0.88,5.06 | | 2.9 | 1.64,5.15 | |
| | O-level/CSE/vocational/none | 3.69 | 1.65,8.25 | | 2.17 | 1.18,3.97 | |
| Interaction terms | Female X A-level | 0.86 | 0.32,2.33 | | 0.86 | 0.43,1.72 | |
| | Female X O-level/etc. | 0.98 | 0.39,2.47 | | 1.32 | 0.64,2.72 | |
| | **P- value for joint test*** | | | 0.6984 | | | 0.157 |
| **Class** | | | | | | | |
| | **Depression only** | | | | | | |
| Main effects | Female | 1.68 | 0.99,2.85 | | 1.98 | 1.01,3.87 | |
| | II | 1.61 | 1.01,2.56 | | 1.59 | 0.84,3.00 | |
| | III | 1.63 | 1.01,2.63 | | 1.84 | 0.93,3.63 | |
| | IV/V | 2.36 | 1.05,5.31 | | 3.77 | 1.27,11.22 | |
| Interaction terms | Female X II | 0.81 | 0.44,1.47 | | 0.6 | 0.29,1.24 | |
| | Female X III | 0.99 | 0.53,1.82 | | 0.66 | 0.30,1.44 | |
| | Female X IV/V | 1.17 | 0.40,3.43 | | 0.29 | 0.06,1.45 | |
| | **Overweight only** | | | | | | |
| Main effects | Female | 0.65 | 0.42,1.02 | | 0.7 | 0.44,1.10 | |
| | II | 0.95 | 0.67,1.33 | | 1.36 | 0.96,1.93 | |

| | | | | | | | |
|---|---|---|---|---|---|---|---|
| | III | 1.15 | 0.81,1.62 | | 1.64 | 1.11,2.43 | |
| | IV/V | 1.06 | 0.55,2.07 | | 1.38 | 0.55,3.46 | |
| Interaction terms | Female X II | 1.44 | 0.87,2.38 | | 1.06 | 0.64,1.77 | |
| | Female X III | 1.51 | 0.91,2.53 | | 1.28 | 0.74,2.22 | |
| | Female X IV/V | 4.82 | 1.96,11.88 | | 2.74 | 0.86,8.67 | |
| | **Comorbidity** | | | | | | |
| Main effects | Female | 3 | 1.00,8.93 | | 1.81 | 0.80,4.10 | |
| | II | 1.43 | 0.48,4.21 | | 1.81 | 0.86,3.79 | |
| | III | 2.67 | 0.94,7.63 | | 2.64 | 1.19,5.84 | |
| | IV/V | 3.01 | 0.68,13.41 | | 4.18 | 1.09,16.07 | |
| Interaction terms | Female X II | 0.93 | 0.28,3.15 | | 0.78 | 0.33,1.87 | |
| | Female X III | 0.86 | 0.26,2.83 | | 0.93 | 0.35,2.42 | |
| | Female X IV/V | 1.89 | 0.34,10.54 | | 0.91 | 0.19,4.34 | |
| | **P- value for joint test*** | | | 0.3025 | | | 0.1428 |
| **Financial difficulties** | | | | | | | |
| | **Depression only** | | | | | | |
| Main effects | Female | 1.38 | 1.11,1.72 | | 1.28 | 0.92,1.79 | |
| | Some | 1.08 | 0.75,1.56 | | 1.31 | 0.80,2.13 | |
| | Fairly difficult | 1.32 | 0.80,2.19 | | 1.53 | 0.73,3.20 | |
| | Very difficult | 1.33 | 0.67,2.66 | | 1.97 | 0.72,5.41 | |
| Interaction terms | Female X Some | 1.17 | 0.74,1.84 | | 1.09 | 0.60,1.97 | |
| | Female X Fairly difficult | 1.5 | 0.80,2.81 | | 0.92 | 0.37,2.28 | |
| | Female X Very difficult | 1.57 | 0.69,3.61 | | 0.63 | 0.16,2.46 | |
| | **Overweight only** | | | | | | |
| Main effects | Female | 0.93 | 0.76,1.13 | | 0.79 | 0.65,0.97 | |
| | Some | 1.21 | 0.90,1.63 | | 1.04 | 0.75,1.43 | |
| | Fairly difficult | 0.83 | 0.52,1.33 | | 0.78 | 0.45,1.35 | |
| | Very difficult | 1.18 | 0.67,2.09 | | 1.44 | 0.72,2.87 | |
| Interaction terms | Female X Some | 0.88 | 0.59,1.32 | | 0.96 | 0.63,1.48 | |
| | Female X Fairly difficult | 2.19 | 1.21,3.99 | | 1.69 | 0.86,3.29 | |
| | Female X Very difficult | 1.11 | 0.49,2.52 | | 1.43 | 0.63,3.29 | |
| | **Comorbidity** | | | | | | |
| Main effects | Female | 2.87 | 1.83,4.50 | | 1.38 | 0.97,1.96 | |
| | Some | 2.36 | 1.26,4.45 | | 0.98 | 0.52,1.86 | |
| | Fairly difficult | 1.76 | 0.69,4.54 | | 0.99 | 0.37,2.67 | |
| | Very difficult | 0 | | | 2.02 | 0.62,6.51 | |
| Interaction terms | Female X Some | 0.65 | 0.31,1.37 | | 1.26 | 0.60,2.65 | |
| | Female X Fairly difficult | 1.15 | 0.37,3.52 | | 2.32 | 0.78,6.86 | |

| | | | | | | | |
|---|---|---|---|---|---|---|---|
| | Female X Very difficult | 0 | | | 1.22 | 0.31,4.80 | |
| | **P- value for joint test*** | | | 0.3065 | | | 0.6584 |

Note: Models adjusted for ethnicity. *Joint test of whether the coefficients on the interaction terms are jointly equal to zero.

**Table 6. Relative indices of inequality for education, class and financial difficulties**

| | Female age 17 | | Female age 24 | | Male age 17 | | Male age 24 | |
|---|---|---|---|---|---|---|---|---|
| **EDUCATION** | | | | | | | | |
| **Depression only** | RR | 95% CI | RR | 95% CI | RR | 95% CI | RR | 95% CI |
| RII | 1.8 | 1.21,2.70 | 1.21 | 0.71,2.07 | 1.83 | 1.05,3.21 | 1.44 | 0.69,3.03 |
| **Overweight only** | | | | | | | | |
| RII | 2.71 | 1.79,4.10 | 3.52 | 2.32,5.34 | 1.88 | 1.22,2.92 | 1.69 | 1.08,2.66 |
| **Both depression and overweight** | | | | | | | | |
| RII | 7.45 | 3.96,14.04 | 4.12 | 2.38,7.11 | 7.35 | 2.40,22.47 | 2.72 | 1.22,6.06 |
| **CLASS** | | | | | | | | |
| **Depression only** | | | | | | | | |
| RII | 1.99 | 1.29,3.07 | 1.46 | 0.83,2.56 | 1.62 | 0.92,2.83 | 2.41 | 1.10,5.29 |
| **Overweight only** | | | | | | | | |
| RII | 2.74 | 1.77,4.23 | 3.07 | 1.99,4.72 | 1.32 | 0.84,2.05 | 1.8 | 1.08,3.00 |
| **Both depression and overweight** | | | | | | | | |
| RII | 4.59 | 2.44,8.62 | 3.9 | 2.09,7.28 | 4.35 | 1.45,13.08 | 3.71 | 1.52,9.07 |
| **FINANCIAL DIFFICULTIES** | | | | | | | | |
| **Depression only** | | | | | | | | |
| RII | 2.7 | 1.66,4.38 | 1.92 | 1.05,3.52 | 1.43 | 0.75,2.73 | 2.26 | 0.96,5.34 |
| **Overweight only** | | | | | | | | |
| RII | 1.74 | 1.06,2.86 | 1.53 | 0.95,2.46 | 1.19 | 0.71,2.02 | 1.03 | 0.56,1.90 |
| **Both depression and overweight** | | | | | | | | |
| RII | 4.21 | 2.15,8.25 | 3.1 | 1.64,5.83 | 3.41 | 1.03,11.28 | 1.32 | 0.45,3.84 |

Note: Models adjusted for ethnicity.

**Table 7. Association between family socioeconomic circumstances and comorbidity at ages 17 and 24 adjusted for additionally for maternal BMI and depressive score**

| | **Female age 17** | | **Female age 24** | | **Male age 17** | | **Male age 24** | |
|---|---|---|---|---|---|---|---|---|
| **EDUCATION** | | | | | | | | |
| **Depression only** | RR | 95% CI | RR | 95% CI | RR | 95% CI | RR | 95% CI |
| Degree | ref | | ref | | ref | | ref | |
| A-level | 1.35 | 1.03,1.78 | 1.38 | 0.99,1.92 | 1.24 | 0.87,1.78 | 1.83 | 1.12,3.01 |
| O-level/CSE/vocational/none | 1.4 | 1.06,1.85 | 1.07 | 0.74,1.55 | 1.44 | 0.99,2.09 | 1.24 | 0.73,2.13 |
| **Overweight only** | | | | | | | | |
| Degree | ref | | ref | | ref | | ref | |
| A-level | 1.13 | 0.84,1.53 | 1.33 | 0.99,1.78 | 1.32 | 0.98,1.77 | 1.5 | 1.10,2.06 |
| O-level/CSE/vocational/none | 1.58 | 1.19,2.12 | 1.89 | 1.42,2.51 | 1.3 | 0.96,1.77 | 1.23 | 0.91,1.68 |
| **Both depression and overweight** | | | | | | | | |
| Degree | ref | | ref | | ref | | ref | |
| A-level | 1.47 | 0.90,2.39 | 2.14 | 1.44,3.19 | 1.84 | 0.76,4.45 | 2.66 | 1.49,4.75 |
| O-level/CSE/vocational/none | 2.67 | 1.68,4.25 | 2.25 | 1.51,3.36 | 3.09 | 1.36,7.02 | 1.8 | 0.97,3.34 |
| **CLASS** | | | | | | | | |
| **Depression only** | | | | | | | | |
| I | ref | | ref | | ref | | ref | |
| II | 1.26 | 0.88,1.82 | 0.92 | 0.61,1.38 | 1.58 | 0.99,2.51 | 1.57 | 0.83,2.98 |
| III | 1.5 | 1.04,2.18 | 1.14 | 0.75,1.74 | 1.56 | 0.96,2.51 | 1.77 | 0.89,3.53 |
| IV/V | 2.42 | 1.19,4.92 | 0.97 | 0.35,2.69 | 2.2 | 0.97,4.99 | 3.58 | 1.17,10.92 |
| **Overweight only** | | | | | | | | |
| I | ref | | ref | | ref | | ref | |
| II | 1.23 | 0.84,1.81 | 1.33 | 0.93,1.90 | 0.85 | 0.60,1.21 | 1.27 | 0.88,1.82 |
| III | 1.48 | 1.00,2.19 | 1.83 | 1.25,2.67 | 0.96 | 0.67,1.38 | 1.44 | 0.96,2.17 |
| IV/V | 3.67 | 1.95,6.90 | 2.65 | 1.25,5.63 | 0.83 | 0.42,1.67 | 1.17 | 0.43,3.15 |
| **Both depression and overweight** | | | | | | | | |
| I | ref | | ref | | ref | | ref | |
| II | 1.15 | 0.64,2.07 | 1.27 | 0.75,2.14 | 1.28 | 0.43,3.79 | 1.67 | 0.79,3.55 |
| III | 1.76 | 0.99,3.13 | 1.99 | 1.16,3.41 | 2.22 | 0.77,6.36 | 2.26 | 1.00,5.08 |
| IV/V | 3.34 | 1.40,7.97 | 2.38 | 0.89,6.36 | 2.38 | 0.53,10.77 | 3.36 | 0.81,13.94 |
| **FINANCIAL DIFFICULTIES** | | | | | | | | |
| **Depression only** | | | | | | | | |
| None reported | ref | | ref | | ref | | ref | |
| Some | 1.18 | 0.88,1.57 | 1.33 | 0.93,1.90 | 1.02 | 0.70,1.47 | 1.24 | 0.75,2.06 |
| Fairly difficult | 1.74 | 1.20,2.54 | 1.25 | 0.76,2.06 | 1.18 | 0.71,1.97 | 1.38 | 0.62,3.07 |
| Very difficult | 1.81 | 1.09,3.01 | 1.08 | 0.44,2.65 | 1.13 | 0.55,2.33 | 1.71 | 0.61,4.76 |
| **Overweight only** | | | | | | | | |
| None reported | ref | | ref | | ref | | ref | |

| | | | | | | | | |
|---|---|---|---|---|---|---|---|---|
| Some | 1.04 | 0.77,1.41 | 0.92 | 0.68,1.23 | 1.14 | 0.84,1.55 | 0.97 | 0.69,1.37 |
| Fairly difficult | 1.81 | 1.20,2.72 | 1.18 | 0.77,1.81 | 0.75 | 0.46,1.24 | 0.72 | 0.40,1.29 |
| Very difficult | 1.28 | 0.70,2.36 | 1.84 | 1.03,3.27 | 1.05 | 0.57,1.92 | 1.34 | 0.64,2.82 |
| **Both depression and overweight** | | | | | | | | |
| None reported | ref | | ref | | ref | | ref | |
| Some | 1.27 | 0.85,1.91 | 1.06 | 0.71,1.58 | 2.2 | 1.15,4.22 | 0.85 | 0.44,1.65 |
| Fairly difficult | 1.53 | 0.85,2.75 | 1.84 | 1.11,3.04 | 1.58 | 0.60,4.21 | 0.77 | 0.27,2.23 |
| Very difficult | 2.51 | 1.30,4.84 | 1.89 | 0.94,3.82 | 0 | | 1.47 | 0.44,4.93 |

Note: Models adjusted for ethnicity.

**Table 8. Association between family socioeconomic circumstances and depression-overweight comorbidity defined by BMI>25 at ages 17 and 24**

| | Female age 17 | | Male age 17 | |
|---|---|---|---|---|
| **EDUCATION** | | | | |
| **Depression only** | RRR | 95% CI | RRR | 95% CI |
| Degree | ref | | ref | |
| A-level | 1.39 | 1.07,1.82 | 1.23 | 0.87,1.75 |
| O-level/CSE/vocational/none | 1.5 | 1.15,1.97 | 1.48 | 1.03,2.12 |
| **Overweight only** | | | | |
| Degree | ref | | ref | |
| A-level | 1.31 | 0.98,1.76 | 1.54 | 1.14,2.07 |
| O-level/CSE/vocational/none | 1.89 | 1.42,2.51 | 1.64 | 1.21,2.22 |
| **Both depression and overweight** | | | | |
| Degree | ref | | ref | |
| A-level | 1.82 | 1.13,2.93 | 2.24 | 0.92,5.44 |
| O-level/CSE/vocational/none | 3.72 | 2.36,5.86 | 3.89 | 1.68,9.03 |
| **CLASS** | | | | |
| **Depression only** | | | | |
| I | ref | | ref | |
| II | 1.29 | 0.90,1.84 | 1.6 | 1.01,2.52 |
| III | 1.62 | 1.13,2.32 | 1.65 | 1.03,2.64 |
| IV/V | 2.69 | 1.33,5.43 | 2.64 | 1.22,5.74 |
| **Overweight only** | | | | |
| I | ref | | ref | |
| II | 1.38 | 0.94,2.04 | 1.02 | 0.72,1.46 |
| III | 1.72 | 1.17,2.55 | 1.24 | 0.87,1.78 |
| IV/V | 5 | 2.72,9.19 | 1.21 | 0.62,2.38 |
| **Both depression and overweight** | | | | |
| I | ref | | ref | |
| II | 1.35 | 0.76,2.39 | 1.34 | 0.46,3.88 |
| III | 2.28 | 1.30,4.00 | 2.61 | 0.92,7.38 |
| IV/V | 5.78 | 2.55,13.12 | 3.28 | 0.73,14.72 |
| **FINANCIAL DIFFICULTIES** | | | | |
| **Depression only** | | | | |
| None reported | ref | | ref | |
| Some | 1.28 | 0.97,1.69 | 1.1 | 0.77,1.57 |
| Fairly difficult | 1.96 | 1.36,2.81 | 1.31 | 0.78,2.20 |
| Very difficult | 1.99 | 1.21,3.28 | 1.5 | 0.77,2.92 |
| **Overweight only** | | | | |
| None reported | ref | | ref | |
| Some | 1.05 | 0.78,1.41 | 1.13 | 0.83,1.54 |
| Fairly difficult | 1.75 | 1.19,2.58 | 0.87 | 0.54,1.39 |
| Very difficult | 1.32 | 0.72,2.40 | 1.27 | 0.72,2.26 |
| **Both depression and overweight** | | | | |

| | | | | |
|---|---|---|---|---|
| None reported | ref | | ref | |
| Some | 1.61 | 1.09,2.36 | 2.22 | 1.17,4.21 |
| Fairly difficult | 2.13 | 1.23,3.69 | 1.64 | 0.63,4.25 |
| Very difficult | 3.64 | 2.00,6.60 | * | |

Note: Models adjusted for ethnicity. * Model did not converge due to low numbers.

**Table 9. Association between family socioeconomic circumstances and depression and obesity comorbidity at ages 17 and 24**

| | **Female age 17** | | **Female age 24** | | **Male age 17** | | **Male age 24** | |
|---|---|---|---|---|---|---|---|---|
| **EDUCATION** | | | | | | | | |
| **Depression only** | RR | 95% CI | RR | 95% CI | RR | 95% CI | RR | 95% CI |
| Degree | ref | | ref | | ref | | ref | |
| A-level | 1.44 | 1.13,1.85 | 1.49 | 1.12,1.97 | 1.25 | 0.89,1.75 | 1.85 | 1.19,2.87 |
| O-level/CSE/vocational/none | 1.62 | 1.26,2.08 | 1.23 | 0.91,1.66 | 1.62 | 1.15,2.28 | 1.42 | 0.91,2.23 |
| **Obesity only** | | | | | | | | |
| Degree | ref | | ref | | ref | | ref | |
| A-level | 2.02 | 1.14,3.59 | 1.7 | 1.08,2.68 | 1.91 | 1.11,3.28 | 2.04 | 1.24,3.35 |
| O-level/CSE/vocational/none | 2.96 | 1.71,5.14 | 3 | 1.98,4.53 | 1.84 | 1.06,3.21 | 1.71 | 1.01,2.91 |
| **Both depression and obesity** | | | | | | | | |
| Degree | ref | | ref | | ref | | ref | |
| A-level | 1.5 | 0.67,3.37 | 2.86 | 1.55,5.29 | 2.88 | 0.57,14.45 | 2.52 | 1.03,6.20 |
| O-level/CSE/vocational/none | 4 | 1.96,8.18 | 3.33 | 1.86,5.94 | 2.8 | 0.52,15.17 | 1.77 | 0.61,5.15 |
| **CLASS** | | | | | | | | |
| **Depression only** | | | | | | | | |
| I | ref | | ref | | ref | | ref | |
| II | 1.24 | 0.89,1.72 | 0.99 | 0.69,1.43 | 1.63 | 1.06,2.52 | 1.56 | 0.91,2.67 |
| III | 1.6 | 1.15,2.24 | 1.32 | 0.91,1.92 | 1.77 | 1.14,2.75 | 1.9 | 1.07,3.37 |
| IV/V | 2.31 | 1.28,4.16 | 1.25 | 0.49,3.14 | 2.51 | 1.17,5.36 | 3.73 | 1.42,9.80 |
| **Obesity only** | | | | | | | | |
| I | ref | | ref | | ref | | ref | |
| II | 1.79 | 0.83,3.85 | 2.06 | 1.11,3.85 | 1.88 | 0.89,3.95 | 1.53 | 0.81,2.88 |
| III | 2.35 | 1.10,5.04 | 3.17 | 1.66,6.06 | 2.05 | 0.96,4.36 | 2.22 | 1.14,4.33 |
| IV/V | 6.9 | 2.65,17.96 | 8.14 | 3.32,19.92 | 3.79 | 1.33,10.77 | 1.94 | 0.52,7.18 |
| **Both depression and obesity** | | | | | | | | |
| I | ref | | ref | | ref | | ref | |
| II | 1.63 | 0.62,4.32 | 1.42 | 0.69,2.93 | 2.02 | 0.23,17.57 | 1.34 | 0.42,4.32 |
| III | 2.46 | 0.94,6.41 | 2.09 | 1.01,4.30 | 2.7 | 0.31,23.47 | 1.8 | 0.51,6.30 |
| IV/V | 5.02 | 1.40,18.04 | 4.15 | 1.38,12.50 | 7.17 | 0.49,104.18 | 3.21 | 0.48,21.46 |
| **FINANCIAL DIFFICULTIES** | | | | | | | | |
| **Depression only** | | | | | | | | |
| None reported | ref | | ref | | ref | | ref | |
| Some | 1.31 | 1.01,1.68 | 1.39 | 1.04,1.87 | 1.16 | 0.84,1.61 | 1.17 | 0.73,1.86 |
| Fairly difficult | 1.79 | 1.28,2.50 | 1.66 | 1.12,2.48 | 1.45 | 0.91,2.33 | 1.42 | 0.71,2.87 |
| Very difficult | 2.27 | 1.46,3.53 | 1.26 | 0.66,2.41 | 1.19 | 0.61,2.33 | 1.89 | 0.85,4.19 |
| **Obesity only** | | | | | | | | |
| None reported | ref | | ref | | ref | | ref | |

| | | | | | | | | |
|---|---|---|---|---|---|---|---|---|
| Some | 1.33 | 0.82,2.13 | 0.85 | 0.55,1.31 | 1.1 | 0.64,1.87 | 1.3 | 0.81,2.07 |
| Fairly difficult | 2.03 | 1.10,3.76 | 1.43 | 0.81,2.53 | 1.06 | 0.49,2.26 | 0.91 | 0.39,2.09 |
| Very difficult | 1.42 | 0.52,3.89 | 1.88 | 1.03,3.43 | 1.45 | 0.59,3.56 | 1.86 | 0.72,4.78 |
| **Both depression and obesity** | | | | | | | | |
| None reported | ref | | ref | | ref | | ref | |
| Some | 1.66 | 0.90,3.05 | 1.03 | 0.61,1.74 | 3.09 | 0.92,10.30 | 1.23 | 0.50,3.04 |
| Fairly difficult | 1.91 | 0.76,4.81 | 1.97 | 1.02,3.81 | 1.32 | 0.15,11.19 | 1.23 | 0.33,4.61 |
| Very difficult | 3.87 | 1.62,9.21 | 2.19 | 1.00,4.81 | * | | 0.81 | 0.00,9.7e+134 |

Note: Models adjusted for ethnicity. * Model did not converge due to low numbers.

**Table 10. Association between family socioeconomic circumstances and depression-overweight comorbidity at ages 17 and 24 in unimputed sample**

| | Female age 17 | | Female age 24 | | Male age 17 | | Male age 24 | |
|---|---|---|---|---|---|---|---|---|
| **EDUCATION** | | | | | | | | |
| **Depression only** | RR | 95% CI | RR | 95% CI | RR | 95% CI | RR | 95% CI |
| Degree | ref | | ref | | ref | | ref | |
| A-level | 1.36 | 1.03,1.80 | 1.45 | 1.00,2.11 | 1.25 | 0.87,1.80 | 1.52 | 0.86,2.68 |
| O-level/CSE/vocational/none | 1.48 | 1.11,1.97 | 1.11 | 0.74,1.68 | 1.45 | 1.00,2.10 | 1.08 | 0.57,2.05 |
| **Overweight only** | | | | | | | | |
| Degree | ref | | ref | | ref | | ref | |
| A-level | 1.32 | 0.98,1.80 | 1.46 | 1.06,2.03 | 1.47 | 1.07,2.01 | 1.67 | 1.13,2.48 |
| O-level/CSE/vocational/none | 1.82 | 1.35,2.46 | 2.07 | 1.50,2.86 | 1.56 | 1.13,2.16 | 1.57 | 1.04,2.37 |
| **Both depression and overweight** | | | | | | | | |
| Degree | ref | | ref | | ref | | ref | |
| A-level | 1.79 | 1.09,2.95 | 2.28 | 1.45,3.58 | 2.22 | 0.90,5.51 | 2.59 | 1.22,5.47 |
| O-level/CSE/vocational/none | 3.71 | 2.33,5.92 | 2.5 | 1.57,3.96 | 4.29 | 1.83,10.09 | 2.31 | 1.05,5.09 |
| **CLASS** | | | | | | | | |
| **Depression only** | | | | | | | | |
| I | ref | | ref | | ref | | ref | |
| II | 1.24 | 0.86,1.79 | 0.89 | 0.56,1.42 | 1.59 | 1.00,2.54 | 1.61 | 0.78,3.31 |
| III | 1.55 | 1.07,2.24 | 1.32 | 0.82,2.14 | 1.48 | 0.91,2.41 | 1.84 | 0.86,3.96 |
| IV/V | 3.21 | 1.56,6.59 | 1.18 | 0.30,4.67 | 2.48 | 1.08,5.70 | 2.82 | 0.48,16.49 |
| **Overweight only** | | | | | | | | |
| I | ref | | ref | | ref | | ref | |
| II | 1.25 | 0.84,1.87 | 1.35 | 0.89,2.04 | 1.04 | 0.72,1.50 | 1.26 | 0.81,1.96 |
| III | 1.69 | 1.13,2.52 | 1.93 | 1.25,2.97 | 1.25 | 0.85,1.83 | 1.33 | 0.82,2.14 |
| IV/V | 6.28 | 3.20,12.33 | 4.57 | 1.80,11.59 | 1.43 | 0.67,3.08 | 3.27 | 1.00,10.68 |
| **Both depression and overweight** | | | | | | | | |
| I | ref | | ref | | ref | | ref | |
| II | 1.24 | 0.68,2.23 | 1.15 | 0.64,2.04 | 1.44 | 0.47,4.40 | 1.65 | 0.68,3.98 |
| III | 2.1 | 1.17,3.75 | 2.12 | 1.19,3.78 | 3.13 | 1.07,9.15 | 1.81 | 0.71,4.60 |
| IV/V | 6.63 | 2.71,16.21 | 3.96 | 1.18,13.27 | 4.75 | 1.01,22.42 | 4.74 | 0.77,29.20 |
| **FINANCIAL DIFFICULTIES** | | | | | | | | |
| **Depression only** | | | | | | | | |
| None reported | ref | | ref | | ref | | ref | |
| Some | 1.22 | 0.91,1.63 | 1.53 | 1.05,2.24 | 1 | 0.68,1.48 | 1.59 | 0.87,2.91 |
| Fairly difficult | 2.07 | 1.41,3.05 | 1.2 | 0.64,2.26 | 1.36 | 0.82,2.26 | 1.57 | 0.64,3.87 |
| Very difficult | 2.14 | 1.26,3.64 | 1.49 | 0.52,4.22 | 1.49 | 0.72,3.08 | 2.15 | 0.55,8.41 |
| **Overweight only** | | | | | | | | |
| None reported | ref | | ref | | ref | | ref | |

| | | | | | | | | |
|---|---|---|---|---|---|---|---|---|
| Some | 1.04 | 0.77,1.42 | 0.9 | 0.64,1.28 | 1.19 | 0.86,1.64 | 1.14 | 0.75,1.74 |
| Fairly difficult | 1.82 | 1.21,2.74 | 1.11 | 0.67,1.85 | 0.83 | 0.49,1.40 | 0.52 | 0.23,1.18 |
| Very difficult | 1.19 | 0.61,2.29 | 2.75 | 1.35,5.63 | 1.57 | 0.84,2.93 | 1.98 | 0.77,5.13 |
| **Both depression and overweight** | | | | | | | | |
| None reported | ref | | ref | | ref | | ref | |
| Some | 1.52 | 1.01,2.30 | 1.16 | 0.73,1.83 | 2.77 | 1.45,5.29 | 0.7 | 0.30,1.64 |
| Fairly difficult | 1.83 | 1.01,3.32 | 2.19 | 1.23,3.89 | 1.93 | 0.71,5.22 | 0.54 | 0.12,2.36 |
| Very difficult | 3.86 | 2.05,7.26 | 4.31 | 1.90,9.80 | * | | 1.69 | 0.35,8.27 |

Note: Models adjusted for ethnicity. * Model did not converge due to low numbers.
